# Temporal evolution of COVID-19 in the states of India using SIQR Model

**DOI:** 10.1101/2020.06.08.20125658

**Authors:** Alok Tiwari

## Abstract

COVID 19 entered during the last week of April 2020 in India has caused 3,546 deaths with 1,13,321 number of reported cases. Indian government has taken many proactive steps, including strict lockdown of the entire nation for more than 50 days, identification of hotspots, app-based tracking of citizens to track infected. This paper investigated the evolution of COVID 19 in five states of India (Maharashtra, UP, Gujrat, Tamil Nadu, and Delhi) from 1^st^ April 2020 to 20^th^ May 2020. Variation of doubling rate and reproduction number (from SIQR) with time is used to analyse the performance of the majorly affected Indian states. It has been determined that Uttar Pradesh is one of the best performers among five states with the doubling rate crossing 18 days as of 20^th^ May. Tamil Nadu has witnessed the second wave of infections during the second week of May. Maharashtra is continuously improving at a steady rate with its doubling rate reaching to 12.67 days. Also these two states are performing below the national average in terms of infection doubling rate. Gujrat and Delhi have reported the doubling rate of 16.42 days and 15.49 days respectively. Comparison of these states has also been performed based on time-dependent reproduction number. Recovery rate of India has reached to 40 % as the day paper is written.

## 1. INTRODUCTION

Coronavirus disease 2019 (COVID-19) originated from the Wuhan city of the Hubei province (China) in December 2019 has caused global pandemic with more than 5 million positive cases and 0.3 million deaths [1]. Most of the countries in the world is affected, and India with the 2^nd^ largest population in the world and 112^th^ ranked medical facility in the world [1] speculated to be one of the severely affected nations of the world [2].

During the initial stages of COVID 19 in India, various mathematical analysis has been done to predict the total number of cases [3], peak prediction [4] as well as on the correlation of the lockdown days on the curve flattening [2]. In my previous work [4], SIQR model is used to quantify the spread of the virus in India using reproduction number (Ro), infection doubling rate and infected to quarantined ratio. Time-dependent transmission parameters like time-dependent reproduction number (Rt) [5] has been proven to be more accurate parameters for the analysis of any infectious disease.

India witnessed its first positive case on 30^th^ January 2020, and as of 21^st^ May, 2020 total positive cases has crossed 1,13,321 with only 3,456 deaths [6]. Strict lockdown of the nation for more than 50 days, mobile application based tracing of infected and sealing off hotspot regions (and other measures) seems to be successfully mitigated the intensity at which spread was predicted.

Contribution of different states of the country to the total reported cases is skewed a lot with states like Maharashtra being one of the severely affected state and state-like Mizoram with just one confirmed positive case till date. State-wise analysis of the growth of cases, instead of the country as a whole, is needed to put forward the precise scenario of COVID 19 in India.

In this paper, a time-dependent analysis of the five major contributing states of India (Maharashtra, UP, Gujrat, Tamil Nadu, and Delhi) is performed. Time-dependent reproduction number using SIQR method and the doubling rate of reported cases has been calculated and analysed. Recovery rate of the country is also reported to check the variation of the recovered population with time.

## 2. ANALYSIS APPROACH

In this paper, two transmissibility parameters are used to analyse the growth of COVID 19 in five major affected states of India. Following part of the section defines the concept of the parameter used:

### 2.1 Time-dependent Reproduction Number (R_t_)

This is one of the widely used parameters used to determine the transmissibility of contagious disease over the time duration (t). It is determined using the Susceptible Infected Quarantine Recovered (SIQR) model used in [4]. SIQR model uses four different rate equations to model the spread of disease defined as follows:

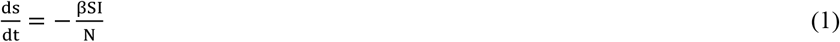

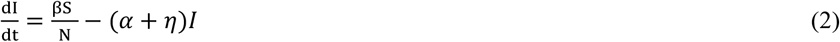

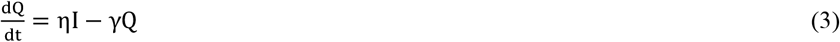

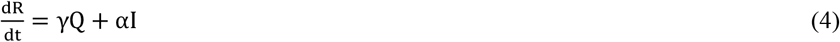

In the following equations, *β* denotes the rate of infection, *η* determines the rate at which new cases are detected from the infected population. *γ* is the rate at which quarantine are getting removed (recovered or died). *α* is the rate of removal of infectious individuals who are asymptotic (or for any reasons) and didn’t get quarantined. Reader can refer to [4] for a better understanding of this model.

Integrating Eq. (2) and addition of Eq. (3) and Eq. (4) gives us the following two equations:

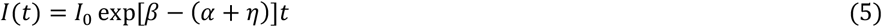

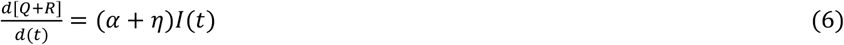

Substitution of Eq. (5) to the Eq. (6) and integration of the resultant over t gives us an equation which is used in the determination of R_t_ :

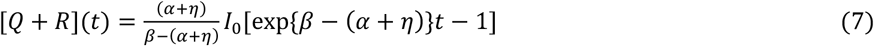

Eq. (7) is fitted with 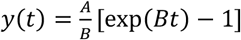 using least square fitting method to determine the coefficient (*α* + *η*) and *β* used in the SIQR model.

Basic reproduction number (R_0_) average over time (t) is defined as the following:

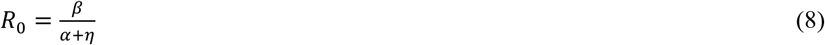

Time-dependent reproduction number (R_t_) in this paper is determined over every fifth day starting 15^th^ April 2020 for each of the five states. Reported positive cases of the states are fitted with Eq. (7) by taking 15 days data for the fitting (Fig. 1).

**Fig 1:**
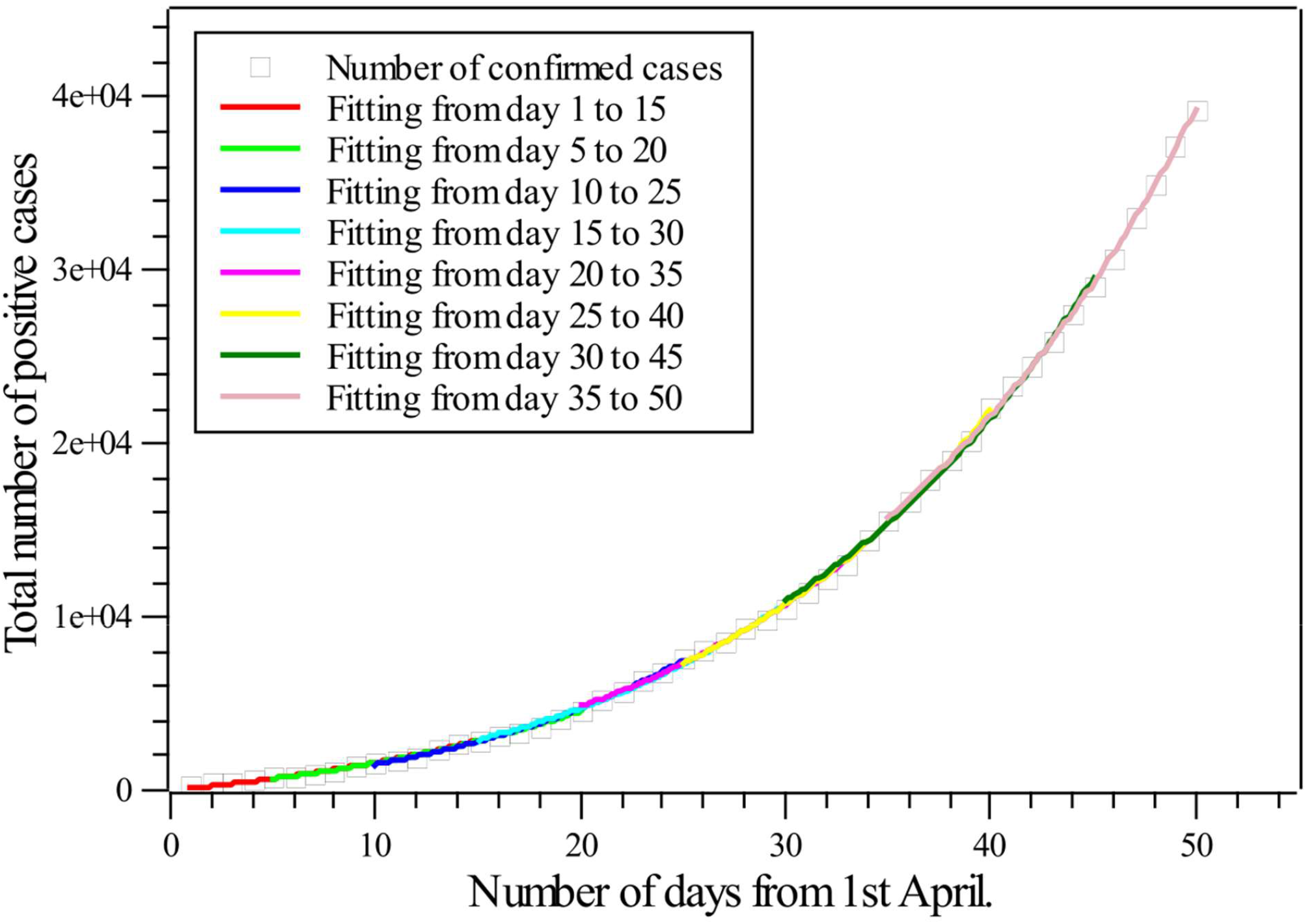
Fitting of Eq. (7) to the reported positive cases of the Maharashtra to get basic reproduction number R_0_ at every fifth day starting from 15^th^ April 2020.

### 2.2 Doubling Rate

This is another widely talked and accepted parameter to quantify the spread of any contagious disease in a region. It is defined as the number of days required for a disease to double its infected population in a region. This number for each of the five states is calculated using the data of total number of reported positive cases from 1^st^ April to 20^th^ May, 2020. The doubling rate for i^th^ day can be defined as follows:

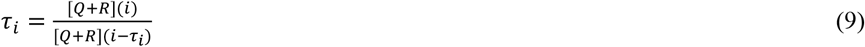

Here *τ*_i_ is the doubling rate at i^th^ day, [Q+R](i) is the total number of reported positive cases on i^th^ day, [Q+R](i-*τ*_i_) is the total number of reported positive cases at day (i-*τ*_i_) when cases are half as compared to i^th^ day.

### 2.3 Recovery Rate

Recovery rate of any contagious disease is defined as the percentage of recovered cases from the total reported positive cases. Recovery rate can be used to compare the quality of health care facility in a particular region or to the immunity of the infected cases. Mathematically, this can be defined as the following.

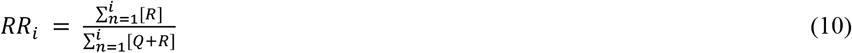

Here, *RR*_i_ is the recovery rate on ith day, 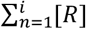 is the total number of recovered individuals in the population on the ith day from day n = 1 and 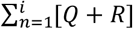 is the total number of reported positive cases on ith day from day n = 1.

## 3. RESULTS AND ANALYSIS

Time-dependent reproduction number and doubling rate discussed in above subsections 2.1 to 2.2 is determined for the major affected states of India using data from [6]. This data is compared with the analysis of the country on whole. Point-wise discussion for each of the five states which contributed to more than 70% of the reported positive cases from the country, is done in the following part of this section.

### 3.1 Maharashtra

Maharashtra (MH) is one of the significantly infected state of the country, and this state contributes 35% to the total reported cases from India. As of 20^th^ May, MH has 39,297 reported positive cases with 1,389 deaths.

With the doubling rate of 5.00 days on 7^th^ April, this state is witnessing continuous but slowly mitigating the wave of spread hit during the first week of April (Fig. 1). MH performance in terms of doubling rate was continuously below the average of the nation. Doubling rate of MH as of 20^th^ May is 12.67 days, lower than other reported four states and equal to of Tamil Nadu (Fig. 1).

If analysed in terms of Rt, MH on 15^th^ April has Rt of 1.17, and on 20^th^ May it downs to 1.12. Even in terms of Rt, MH is continuously improving but at a very slow rate, which is lower than the national average rate (Fig. 2).

**Fig 2:**
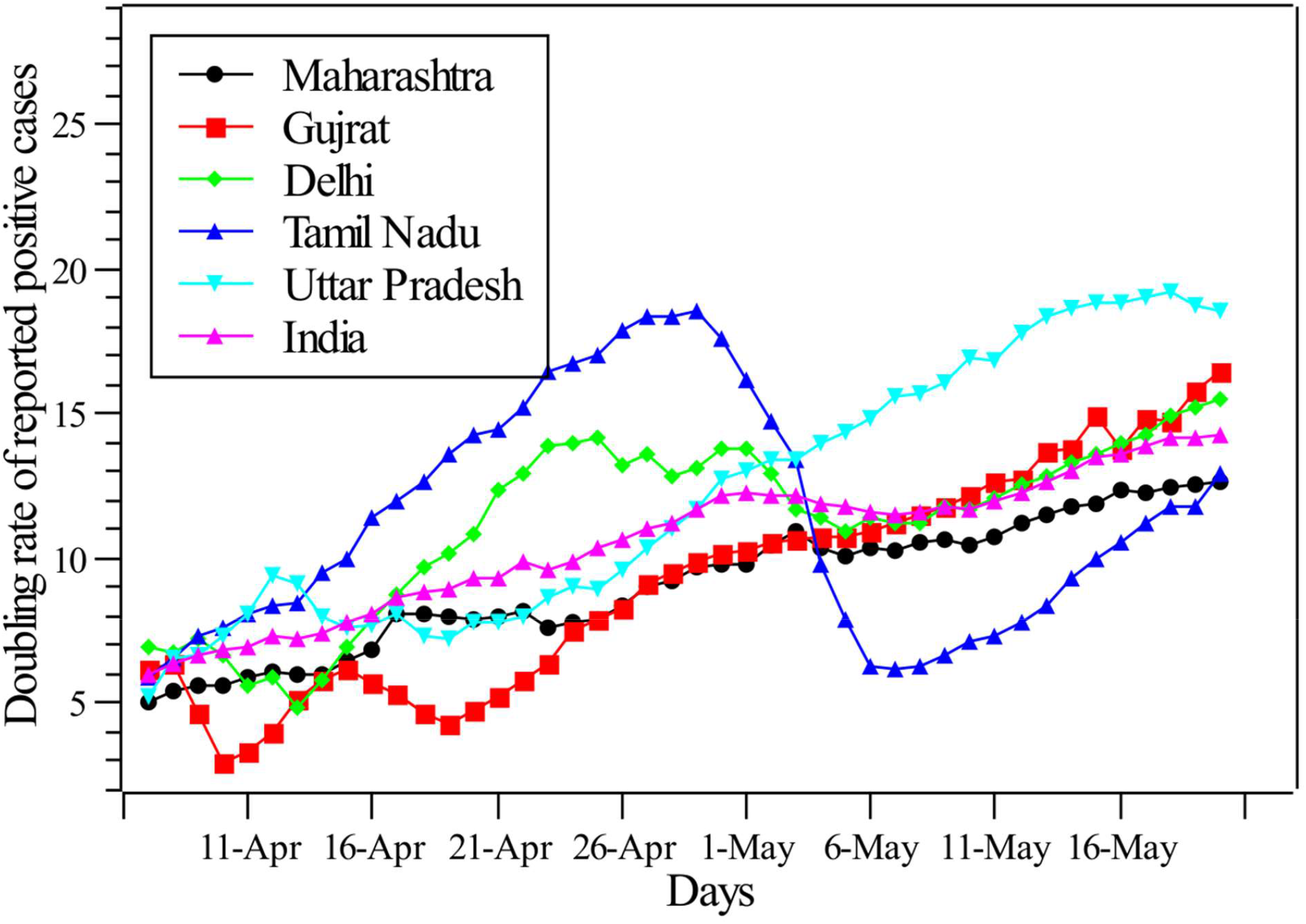
Doubling rate of the reported positive cases for each of the five states and the country as well from 7^th^ April – 20^th^ May 2020.

#### 3.2 Gujrat

Gujrat in terms of the doubling rate at the start of the epidemic (15^th^ April) has the doubling rate of 6.20 days better than all the other four states included in the study (almost same of Delhi). Gujrat witnessed the two dip in doubling rate during the first week and third week of April. However, Gujrat is improving continuously from the month of May, making Gujrat to reach a doubling rate of 16.42 days as of 20^th^ May better than the national average (Fig. 2).

In terms of Rt, Gujrat on 15^th^ April has Rt of 1.56 and reaching its maximum of 2.12 during the second week of April followed by surge during the rest of the month. It has reached to 1.03 on 20^th^ May (Fig. 3).

**Fig 3:**
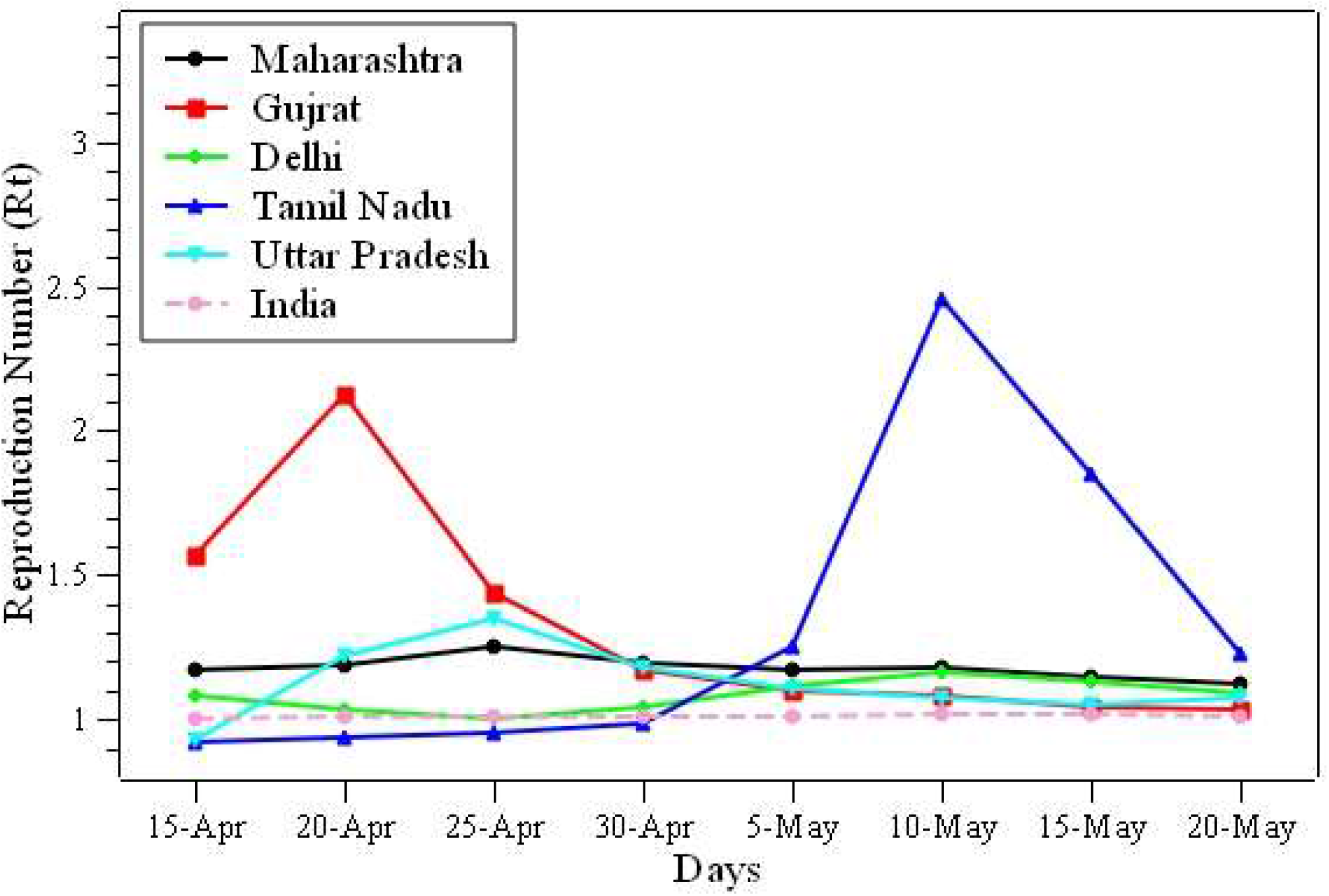
Time dependent reproduction number of all the five states with the country’s average from 15^th^ April to 20^th^ May 2020.

#### 3.3 Delhi

Delhi is the best performer during initial days, started with a doubling rate of 6.92 days followed by dip during the second week of April and the first week of May. This state as of 20^th^ May has the doubling rate of 15.49 days, higher than the national average (Fig. 2).

In terms of Rt, Delhi started with 1.08 on 15^th^ April with a slight dip during the second week of April and followed by steady improvement in Rt. As of 20^th^ May Rt is 1.08, which is higher than the national average of 1.01 (Fig. 3).

#### 3.4 Tamil Nadu

This state witnessed many fluctuations in the number of reported positive cases, started with the doubling rate of 5.85 days, followed by a very high doubling rate of 18.53 during the end of April. Tamil Nadu witnessed the second wave of infection during the first and second week of May when the doubling rate gets down to 6.17 days (very less than the nation’s average). As of 20^th^ May, this state has improved quite significantly, making its doubling rate to reach 12.92 days (lower than the country’s average (Fig. 2).

Recovery rate of the country as discussed in section 2.3, is defined as the ratio of the total number of recovered population to the total reported positive cases. From recovery rate of 12% on 15^th^ April, India has tremendously improved it to reach 40.54 % as of 20^th^ May (Fig. 4). This much percentage of recovered positive cases somewhat indicates the flattening of the peak with the active number of cases getting constant.

**Fig 4:**
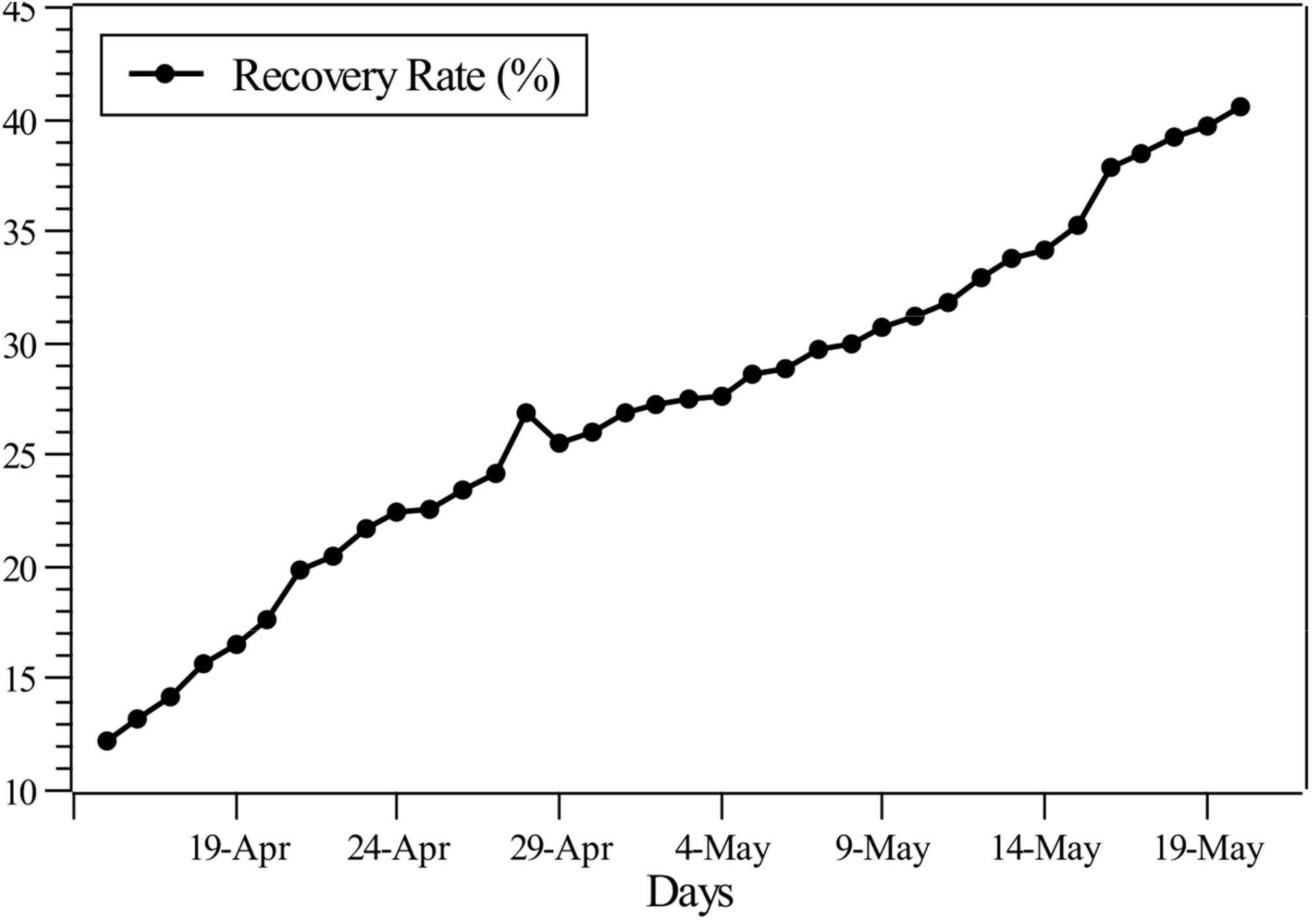
Recovery rate of India from 15^th^ April to 20^th^ May 2020.

In terms of time-dependent reproduction number, this state has witnessed whooping Rt of 2.45 during the second week of May, followed by a drastic improvement, making it to reach 1.22 (Fig. 2).

#### 3.5 Uttar Pradesh

Uttar Pradesh, one of the biggest populated state of the country, has performed unexpectedly well during this epidemic. Started with doubling rate of 5.2 days one of the lowest (better than MH) is improving continuously with better performance than national’s average. As of 20^th^ May, this state has 18.53 days highest in comparison to all other four states (Fig. 1).

In terms of Rt, this state has got a slight increase during the last week of April but has improved quite well, and as of 20^th^ May Rt is 1.07 (Fig. 2).

## 4 CONCLUSION

In this paper, the reproduction number and doubling rate of five major affected states of India are analysed using time-dependent reproduction number and doubling rate. Time-dependent reproduction number is determined using SIQR model. Following points conclude the whole state-wise analysis –

i. Uttar Pradesh is one of the best performers during this epidemic in comparison to four other analysed states.
ii. Tamil Nadu has witnessed major fluctuation in this pandemic with cases rising up again during the second week of May.
iii. Maharashtra and Tamil Nadu are performing below the national average in terms of doubling rate.
iv. Reproduction number of all the states are closed to the national average as of 20^th^ May 2020.
v. Recovery of India is increasing continuously making it to 40% of the total reported positive cases.

## Data Availability

All data used in this available over the web

https://www.covid19india.org/

